# Estimating the impact of test-trace-isolate-quarantine systems on SARS-CoV-2 transmission in Australia

**DOI:** 10.1101/2023.01.10.23284209

**Authors:** Freya M. Shearer, James M. McCaw, Gerard Ryan, Tianxiao Hao, Nicholas J Tierney, Michael Lydeamore, Kate Ward, Sally Ellis, James Wood, Jodie McVernon, Nick Golding

## Abstract

We report on an analysis of Australian COVID-19 case data to estimate the impact of TTIQ systems on SARS-CoV-2 transmission in 2020–21. We estimate that in a low prevalence period in the state of New South Wales (tens of cases per day), TTIQ contributed to a 54% reduction in transmission. In a higher prevalence period in the state of Victoria (hundreds of cases per day), TTIQ contributed to a 42% reduction in transmission. Our results also suggest that case-initiated contact tracing can support timely quarantine in times of system stress. Contact tracing systems for COVID-19 in Australia were highly effective and adaptable in supporting the national suppression strategy through 2020 and 2021.

## INTRODUCTION

Test-trace-isolate-quarantine (TTIQ) strategies have been widely employed to mitigate the impacts of SARS-CoV-2, particularly during the pre-vaccine era of the pandemic. The aim of TTIQ is to reduce transmission through the timely detection and isolation of cases. The potential utility of TTIQ measures is pathogen-specific, as it depends on inherent pathogen characteristics, including transmissibility, and the extent of pre-symptomatic and asymptomatic transmission [1, 2]. Early in 2020, evidence of significant pre-symptomatic [3] and asymptomatic transmission of SARS-CoV-2 [4] indicated that symptomatic case isolation alone, while important for mitigating transmission, was unlikely to be an effective containment strategy. However, estimates of around four days or longer for the generation interval of SARS-CoV-2 [5, 6, 7] (the time between an individual becoming infected and then infecting another person) was considered to be sufficiently long for contact tracers to identify contacts before they became infectious, suggesting the potential for contact tracing and quarantine to have a large additional impact on transmission, provided that tracing was rapid and reasonably complete. Indeed, early modelling studies anticipated an important role for TTIQ in mitigating transmission, though unless a high proportion of cases could be isolated and contacts rapidly traced and quarantined, wider social measures would also be required to achieve outbreak control [8, 9, 10].

A range of contact tracing strategies are possible and approaches to TTIQ for SARS-CoV-2 have varied across settings and through the course of the pandemic. The identification and notification of contacts is generally a manual process managed by public health authorities. Faster and more complete contact identification is theoretically possible with smartphone-based digital contact tracing[11, 12], and while digital approaches were employed in many countries to augment more conventional processes, implementation issues have largely prevented them from contributing meaningfully to disease control [13, 14]. During the 2020–21 (pre-Omicron) period, standard procedure in Australia was to identify contacts via telephone interviews with confirmed cases. Contacts were then directed to quarantine by a public health official. This procedure is highly effective at reducing transmission but the per case burden on public health response teams is high, and systems can rapidly become overwhelmed at high caseloads. When caseloads exceed public health workload capacity, delays from case notification to interview and from interview to contact quarantine increase, reducing TTIQ effectiveness [9, 15].

In countries pursuing a ‘strong suppression’ strategy, including Australia through 2020 and much of 2021 [16], high performing TTIQ systems supported virus suppression and low daily case incidence was critical to the ongoing effectiveness of these systems. In 2020 and 2021, Australia’s most populous state of New South Wales (NSW) effectively managed a series of small outbreaks (tens of cases per day) of COVID-19 through a combination of intensive case and contact management and limited wider social restrictions (with strict border measures also in place). Contact tracing was both upstream for cases with no known source of infection, identifying individuals who had contact with the case during the time in which the case was likely to have acquired their infection, and downstream, identifying individuals who had contact with a confirmed case during the time in which the case was likely to be contagious. Furthermore, tracing of ‘secondary contacts’, *i*.*e*., identifying contacts of contacts, was performed when capacity allowed [17, 18]. Weekly reporting on the performance of NSW contact tracing systems for the latter half of 2020 indicated that 100% of cases were interviewed within 24 hours of notification and 100% of close contacts identified by cases were directed to quarantine by public health officials within 48 hours of case notification [19]. Formal analysis of various case and behavioural data streams during this period also suggest that TTIQ played a key role in outbreak control [20].

The availability of COVID-19 vaccines in early 2021 triggered a transition from a national goal in Australia of no community transmission to one of manageable transmission (from a public health and clinical perspective) [21]. This transition recognised that under higher caseloads, new approaches to contact tracing were required — which included ‘case-initiated’ contact tracing, whereby cases are instructed to self-identify their primary close contacts and ask them to quarantine, and seek a test. In the (relatively) early stages of vaccine roll-out, an incursion of the Delta variant of SARS-CoV-2 in June 2021 led to the largest epidemic wave in Australia (up to thousands of cases per day)[22] prior to the emergence of Omicron in late 2021. To adapt to higher caseloads than previously experienced in NSW, local health authorities adopted a policy of case-initiated contact tracing in August 2021. This included instantaneous result notification via text message to cases, simultaneously directing them to isolate and notify their close contacts.

Here we report on an analysis of Australian COVID-19 case data to estimate the effectiveness of TTIQ systems in 2020-21. First, we directly estimate from detailed case data the contribution of TTIQ to transmission reduction in situations of low prevalence — tens of cases per day or hundreds of cases per day. We then apply a simulation approach, validated by case data, to assess the impact of case-initiated contact tracing on TTIQ effectiveness in NSW during the Delta epidemic from July–November 2021.

## METHODS

### Setting

We focus on the public health response in two of Australia’s eight states/territories: New South Wales (NSW) and Victoria (VIC). NSW and VIC are Australia’s two most populous states, with populations of more than 8 million and 6 million, respectively, largely concentrated in major urban centres. They also have the highest volumes of international travellers and experienced considerably higher total numbers of confirmed COVID-19 cases and peak daily incidence than other jurisdictions in 2020–21. Australia’s adoption of a ‘strong suppression’ strategy in early 2020 meant that COVID-19 epidemiology through much of 2020 and 2021 was characterised by periods of zero case incidence and sporadic outbreaks, as detailed elsewhere [23]. However, a number of major waves of infection occurred, most notably a wave of ancestral virus infection in VIC from June–October 2020 and waves of the Delta variant seeded into NSW and VIC in mid-2021 (Figure 1).

**Figure 1.**
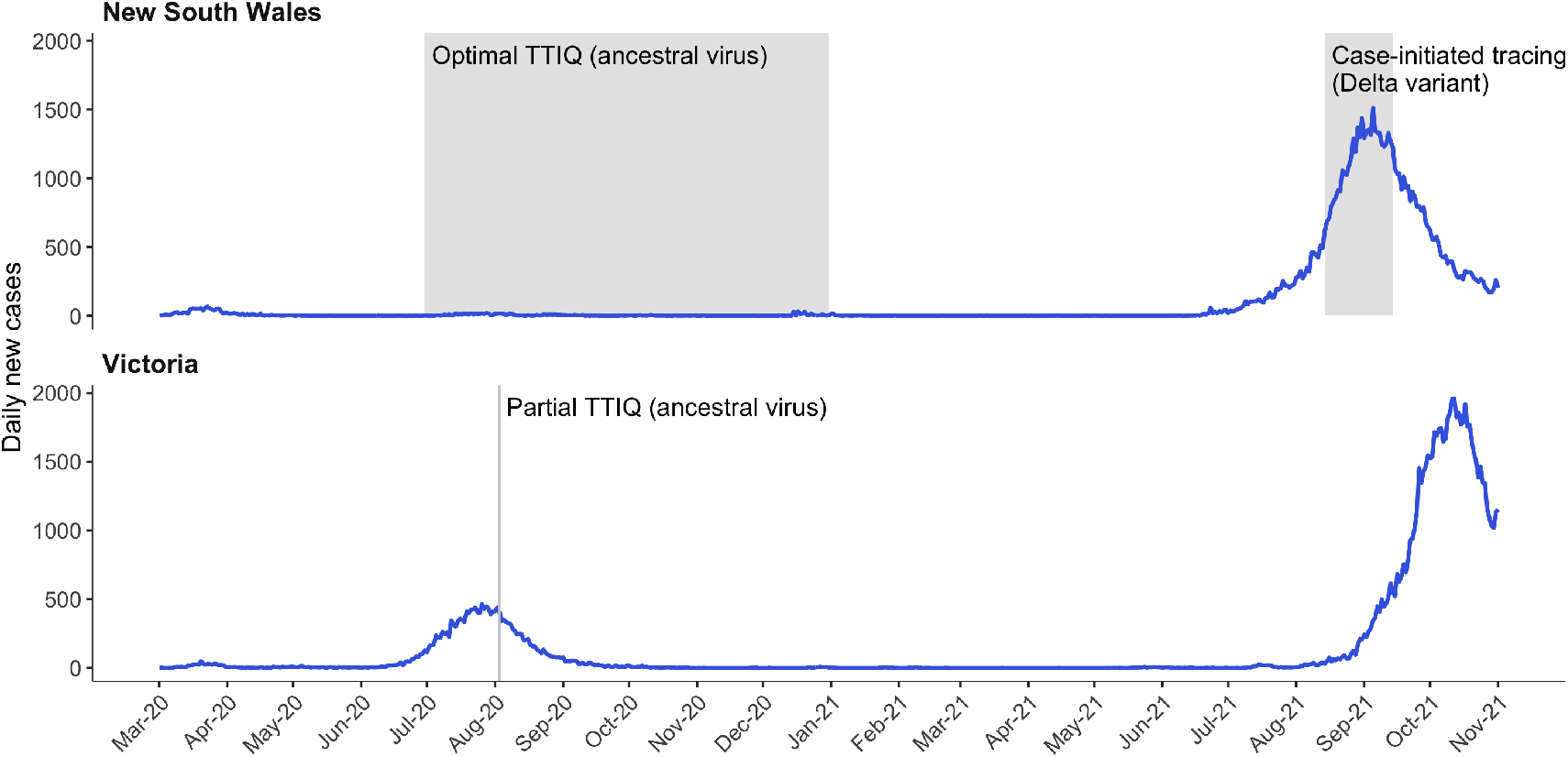
Daily new cases of COVID-19 reported in the states of New South Wales and Victoria from 1 March 2020 to 1 November 20201. Grey shading indicates the time periods/points of case data from which Optimal (tens of cases per day), Partial (hundreds of cases per day), and NSW Delta (where a policy of case-initiated contact tracing was in place, and thousands of cases were reported per day) TTIQ effects were calculated. Details on the response context during each of these periods are provided in Table 1.

### Case data

We use line listed data on cases from NSW and VIC. These data include the dates of key events in the TTIQ pathway: date of test, date of notification (of a positive test result to the public health authority), date of symptom onset (where applicable), and date of case interview. Additionally, the NSW data contain the date of likely infection/exposure and the date of case isolation. Note that for cases detected via contact tracing, the date of isolation can be prior to the date of test (*i*.*e*., it is technically the date of either *quarantine* or isolation).

**Table 1.**
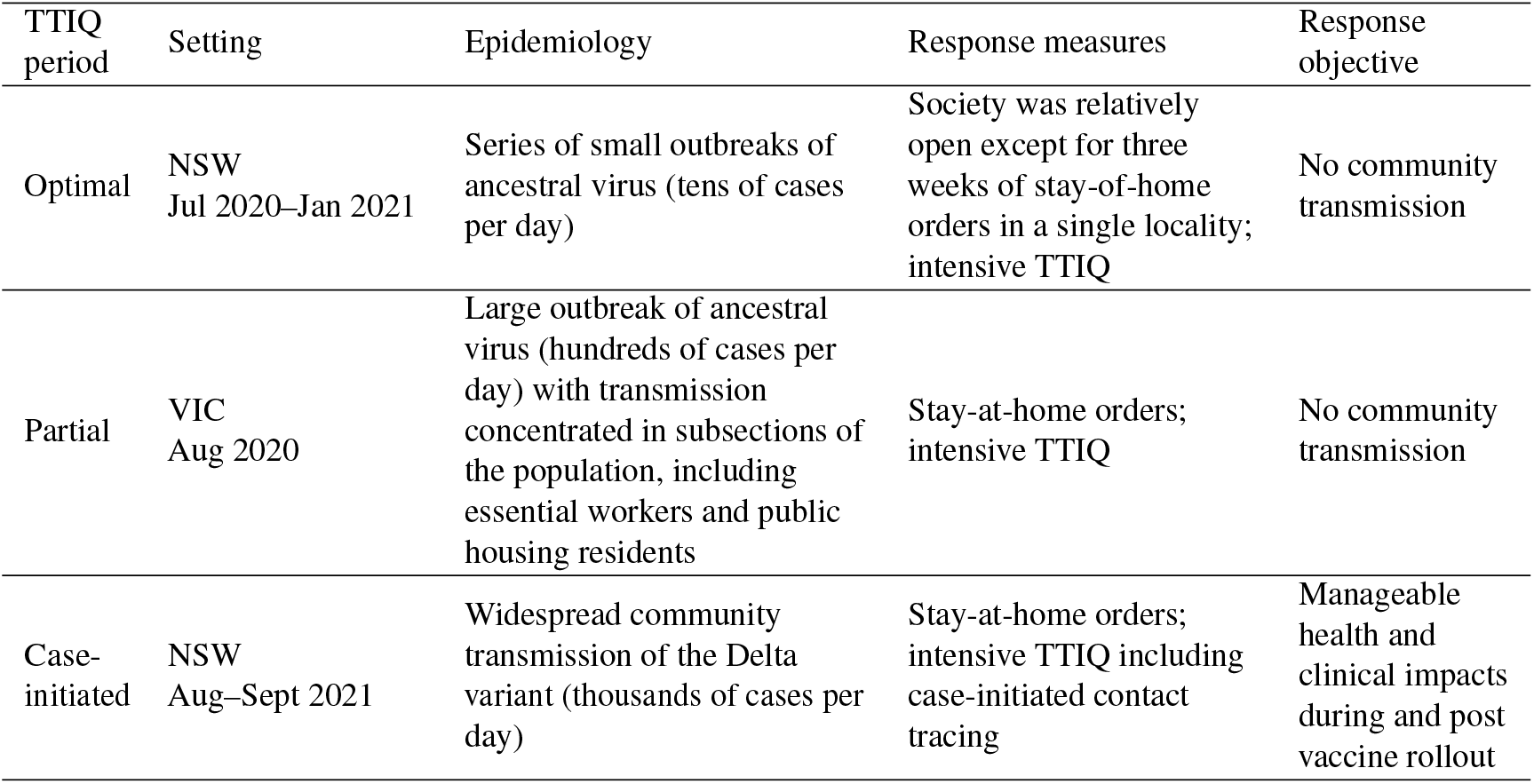
Location and dates of each TTIQ period and the associated epidemiological context, key response measures, and the response objective.

| TTIQ period | Setting | Epidemiology | Response measures | Response objective |
| --- | --- | --- | --- | --- |
| Optimal | NSW<br>Jul 2020–Jan 2021 | Series of small outbreaks of ancestral virus (tens of cases per day) | Society was relatively open except for three weeks of stay-of-home orders in a single locality; intensive TTIQ | No community transmission |
| Partial | VIC<br>Aug 2020 | Large outbreak of ancestral virus (hundreds of cases per day) with transmission concentrated in subsections of the population, including essential workers and public housing residents | Stay-at-home orders; intensive TTIQ | No community transmission |
| Case-initiated | NSW<br>Aug–Sept 2021 | Widespread community transmission of the Delta variant (thousands of cases per day) | Stay-at-home orders; intensive TTIQ including case-initiated contact tracing | Manageable health and clinical impacts during and post vaccine rollout |

### Estimating impact of test-trace-isolate-quarantine systems from case data

We estimate the impact of TTIQ on transmission for two epidemiological and response contexts within the ‘strong suppression’ policy era: very low case loads (tens of cases per day), representing a ‘Optimal’ TTIQ effect; and relatively higher case loads (hundreds of cases per day) representing a ‘Partial’ TTIQ effect (Figure 1 and Table 1). We evaluate the impact of TTIQ in each context by computing a change in expected onward transmission.

The impact of TTIQ systems on transmission depends on both how quickly cases are found and isolated, and what proportion of infections are detected (since undetected infections are unlikely to isolate). Our analyses assume perfect detection of infections and compliance with isolation. While data to estimate case ascertainment in Australia throughout 2020–21, such as from infection prevalence studies, do not exist, ascertainment of infections was likely very high during the ‘strong suppression’ era. This was due to the intensive contact tracing and epidemiological investigation, with the explicit aim of identifying all infections in chains of transmission. Further, Australian jurisdictions repeatedly returned to periods of zero case incidence following outbreaks, suggesting negligible undetected transmission.

#### ‘Optimal’ TTIQ effect

We estimate the effectiveness of TTIQ at reducing transmission using the empirical distribution of times from infection to isolation for all cases in NSW between July 2020 and January 2021. During this period, intensive upstream and downstream contact tracing was carried out, caseloads were low (tens per day), and the impact of TTIQ was clear in suppressing transmission [19]. While some social restrictions were in place, including the closure of nightclubs and person density rules for public venues and workplaces, society remained relatively open. In mid-December 2020, two large super-spreading events occurred in the NSW locality of Northern Beaches, leading to a rapidly spreading outbreak and imposition of stay-at-home orders in the affected locality for three weeks, in addition to an intensive TTIQ response. The outbreak was largely confined to this single locality and never exceeded 70 cases per day.

We directly translate these intervals from infection to isolation into reductions in potential for onward transmission, as described below. We assume that infectiousness over time since infection follows a lognormal distribution (as estimated by [5]) and evaluate the overall percentage reduction in infectiousness due to TTIQ. We define this reduction as a Optimal TTIQ effect, representing the maximum reduction in transmission that we may expect due to TTIQ for that pathogen (*i*.*e*., ancestral virus strain), given the policy, social, and health system context.

We calculate the expected reduction in transmission (*s*; a multiplier on the reproduction number) due to isolation as the finite sum (up to a maximum of *N* = 20 days) of the product of the probability mass functions of the isolation delay distribution (*π*_*I*_, delay from infection to isolation) and generation interval distribution in the absence of isolation (*π*_*G*_, delay from infection to onward transmission for individuals not in isolation):

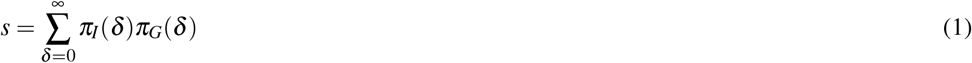

Since the data are only available on a daily time step, we model both *π*_*I*_ and *π*_*G*_ as discrete probability distributions over times since infection to isolation. The generation interval distribution is modelled as a discretised log-normal, with parameters given posterior mean estimates from [5]:

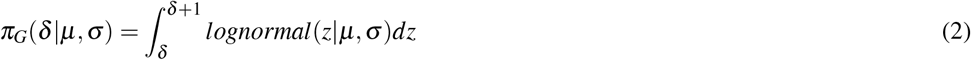

When cases are rapidly found and placed into isolation, the isolation delay distribution, *π*_*I*_, has most of its mass on small delays and the isolation effect *s* tends toward 0. At times when cases are not found and isolated until after most of their infectious period has passed, *π*_*I*_ has most of its mass on large delays and *s* tends toward 1.

#### ‘Partial’ TTIQ effect

Similarly detailed data on the date of isolation of cases were not available from VIC, where a major wave of infection (hundreds of cases per day) occurred from July–October 2020. As in NSW, intensive upstream and downstream contact tracing was in place, and with relatively higher caseloads, contact tracing systems were reported to be under stress [24]. Australia’s Common Operating Picture reported that 10% of case interviews in VIC were outstanding 48 hours after case notification and public health workforce status, including surge capacity, was listed as “saturated” on 27 August 2020 [24]. Stringent stay-at-home orders were in place at this time, and while these measures reduced average contact rates at the population-level, transmission was concentrated in subsections of the population with higher-than-average rates of contact (*e*.*g*., health care workers) [20]. Nonetheless, contacts would have been more easily identifiable than in a freely mixing population, with most contacts occurring in household and workplace settings.

By assuming improvements in TTIQ are proportional to improvements in times to detection (*i*.*e*., times from symptom onset to test), we estimated the relative performance of TTIQ during this period/epidemiological context where TTIQ was considered to be less effective. More specifically, we estimated a distribution of times to isolation using the case data for VIC from 4 August 2020 — the peak of daily locally-acquired COVID-19 cases in Australia in 2020. We define the resulting estimated reduction in transmission as a Partial TTIQ effect.

We interpolated the distribution of times from infection to isolation at each date *t* in VIC, by first defining a time-series of weights *w*(*t*) based on improvements over time in the delay from symptom onset to notification. That is, we applied equation 1 to the distribution of times from *assumed infection* (taken to be 5 days prior to the recorded date of symptom onset [ref]) to *notification*, 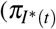 at each time *t*, took the reciprocal of this (so that greater reductions in transmission correspond to higher weights), and normalised it to have maximum value 1:

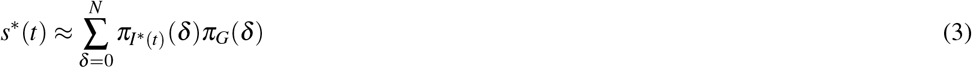

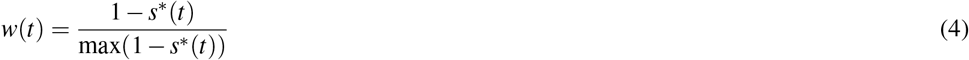

These weights were used to compute an interpolated time-series of distributions of times from infection to isolation 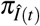 between the time-varying distribution of times from assumed infection to notification 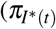, which can be considered an upper bound on the time from infection to isolation), and the Optimal period distribution of times from infection to isolation 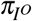 (derived from the NSW case data) as follows:

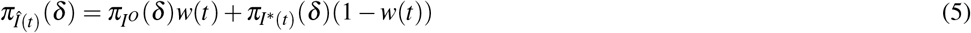

This assumes that when operating at its best (the shortest delays from symptom onset to notification), the Victorian contact tracing system was operating with the same distribution of times from infection to isolation as in NSW during the Optimal reference period. It also assumes that reductions in the distribution times from infection to isolation (or assumed infection to notification) scaled according to improvements in the times from symptom onset to notification.

Finally, we took the infection to isolation distribution for the Partial TTIQ period 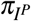 to be the value of the interpolated distribution of times from infection to isolation 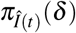 as on 4 August 2020 in VIC.

### Assessing impact of case-initiated contact tracing via simulation

We use a simulation approach to consider the impact of case-initiated contact tracing on transmission in NSW in mid-2021, during the escalating phase of the Delta epidemic wave where caseloads (thousands per day with a peak of 1,511 cases per day) were unprecedented in the Australian context. During this period, intensive upstream and downstream TTIQ were performed, including case-initiated tracing of downstream contacts, and stay-at-home restrictions were in place. We validate our simulations by relating them to available data on timeliness of case isolation from NSW over various time periods (Figure 1).

#### Model overview

A stochastic simulation model was used to represent the relationship between contact tracing delays, symptomatic detection, and times from infection to isolation in continuous chains of contact tracing. By sampling from distributions of contact tracing times, this model generates distributions of time from infection to isolation, which were then used to calculate expected reductions in transmission. Note that the model assumes perfect detection of infections and compliance with isolation.

We calculate the expected distribution of times from infection to isolation (*T*_*I*_) from two different delay distributions: the time from infection of the source case to infection of the contact (*T*_*G*_, the generation interval); and a random sample from the distribution of times from isolation of the source case to isolation of the infected contact (*T*_*A*_, the contact tracing delay) (Figure 2). The latter of these is comprised of several different component distributions, including the time from swab collection to case notification, the time from case notification to case interview, and the times from interview to contact notification and swab collection (Figure 3).

**Figure 2.**
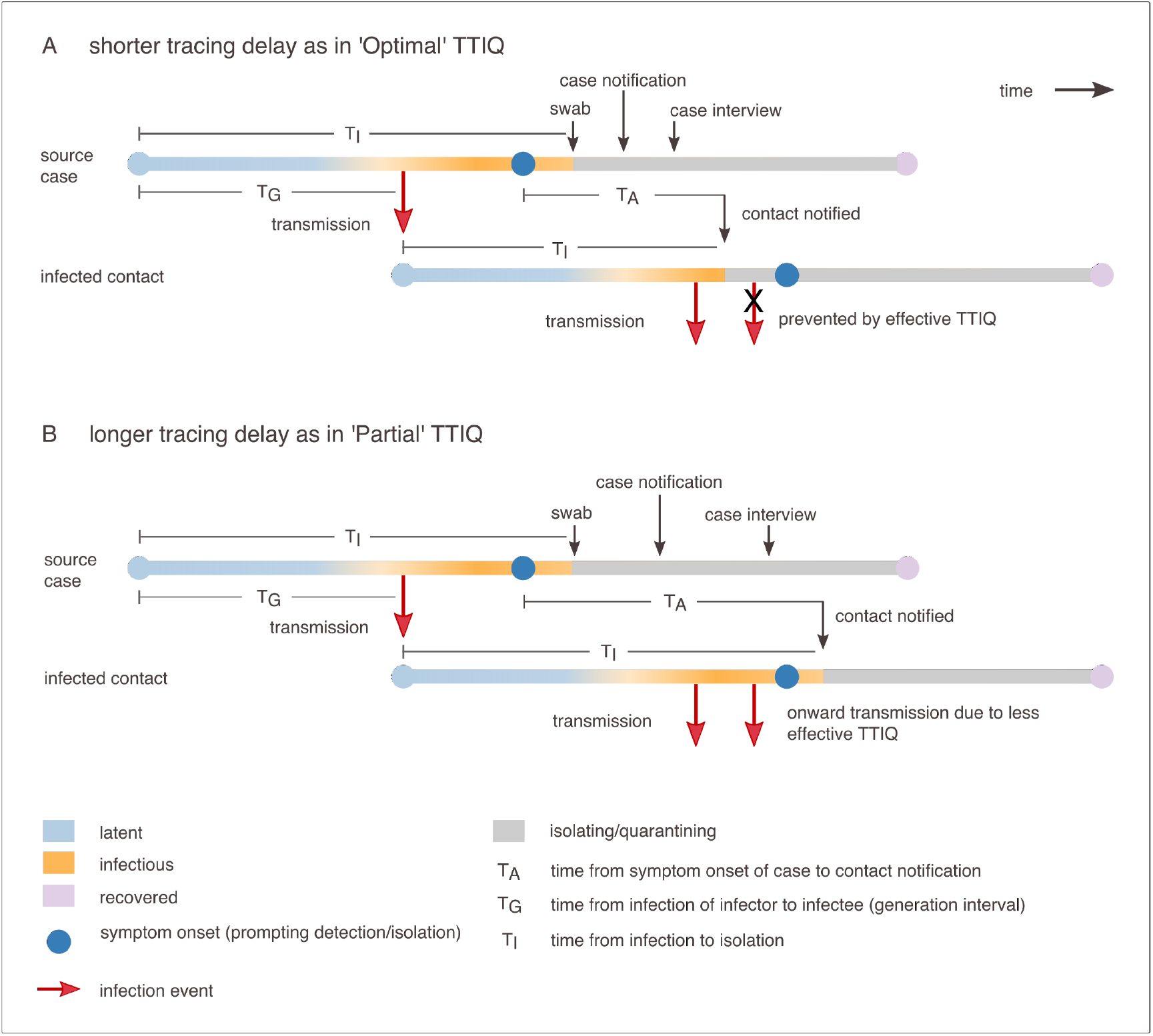
Representation of infection progression for a source case and an infected contact, with the timings of key TTIQ actions indicated. TTIQ aims to reduce the time from infection to isolation (*I*_*A*_). Longer tracing delays (*T*_*A*_) result in longer times to quarantine/isolation and a greater proportion of infectiousness in the community (orange) compared to when delays are short.

**Figure 3.**
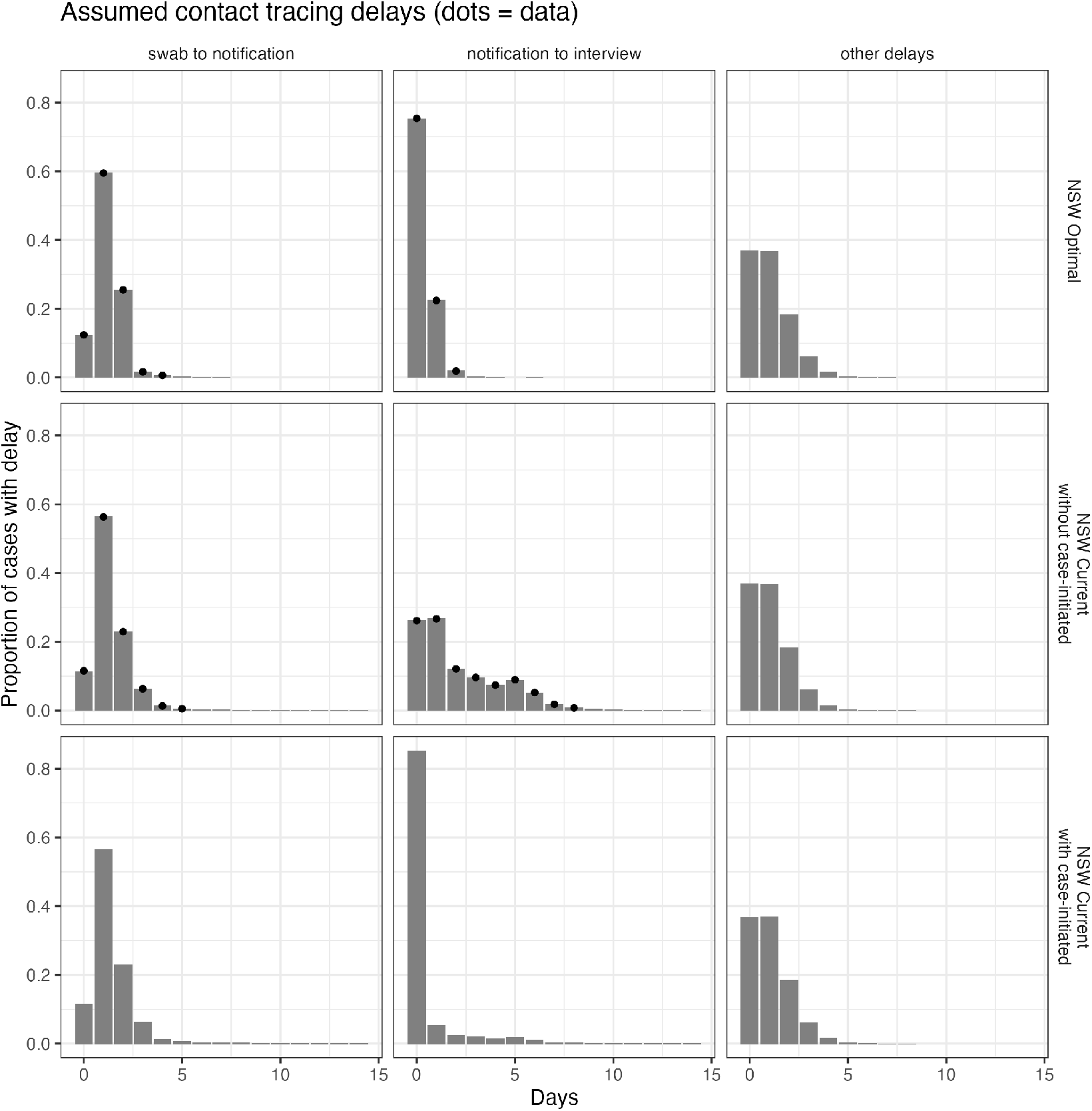
Modelled distributions of various delays in the contact tracing process as estimated from NSW data (dots = data). These distributions are used as inputs in our model of TTIQ impact on transmission. Time from swab collection to notification and notification to interview are informed by NSW data from July 2020 to February 2021 (‘NSW Optimal’, row 1) and from mid-August to mid-September 2021 (‘NSW Delta epidemic without case-initiated’, row 2). ‘Other delays’ is calibrated to match the overall distribution of delays from infection to isolation for the ‘Optimal’ period and has a mean delay of one day. This represents all other delays in the contact tracing process that we are not yet to estimate from data. For example, the time from interview to contact notification and the time from contact notification to isolation. ‘NSW Delta epidemic with case-initiated’ (row 3) assumes the same delays as for ‘NSW Delta epidemic without case-initiated’ except that 80% of notification to interview delays are set to zero. This represents a high proportion of contacts being immediately advised by the case to isolate (*e*.*g*., household contacts).

We estimate the distribution over times from infection to isolation (*T*_*I*_) by iteratively sampling infector and infectee times to isolation and times to onward transmission in long simulated chains of transmission via a recursive sampling algorithm (Markov chain Monte Carlo using a Gibbs sampler) whereby each infectee becomes the infector for the next iteration. This yields a stable distribution of times from infection to isolation for cases from which the reduction in transmission potential can be calculated.

The Gibbs sampling algorithm proceeds as follows, and encodes the schematic in Figure 2. We initialise the Markov chain by sampling an initial time from infection to isolation *T*_*I*_ for an index case. This initial value is rapidly ‘forgotten’ by the chain as it moves to its stable state, and therefore results are insensitive to the choice of distributions from which to sample initial values. We sampled the initial value from a very broad positive-truncated normal distribution as is standard practice for use of Gibbs samplers in Markov chain Monte Carlo sampling in Bayesian inference.

We then iterate the Gibbs sample as follows, incrementing the iteration variable *i* until a predetermined number of samples have been obtained:

1. Sample a generation interval 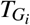 from the infectee to the infector from the generation interval distribution *π*_*G*_, truncated so that the infection cannot occur after the infector’s date of isolation 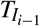:

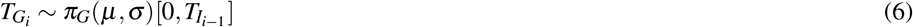
2. Sample a contact tracing delay between detections of the infector and the infectee 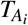 from the contact tracing delay distribution, independently of the other parameters in the Gibbs sampler. The contact tracing delay distribution is parameterised as a categorical distribution with the vector *p*_*F*_ of probability masses (the frequency of delays) for each possible number of days delayed:

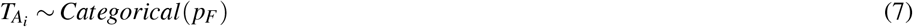
3. Calculate the isolation delay for the infectee 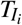 from these other delays. The time from the infector being infected to the infectee being isolated is 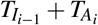, and subtracting the generation interval from this gives the isolation delay for the infectee:

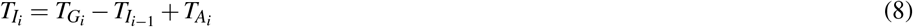
4. Increment *i* so that the infectee becomes the infector for the next iteration of the Gibbs sampler.

This Gibbs sampler yields (non-independent) samples from 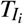, from which the discrete distribution *π*_*G*_ can be calculated, and used in Equation 1

Within this modelling framework, we can investigate the likely impacts of various proposed TTIQ strategies by adjusting parameters and distributions to represent the implementation of those strategies.

#### Modelling case-initiated contact tracing

We model scenarios of TTIQ with and without case-initiated contact tracing by modifying the overall contact tracing delay: the distribution of times from source case isolation to infected contact isolation.

We model the contact tracing delay as the sum of three other types of delay: the time from swab collection to notification, the time from notification to the infected contact being identified (via interview or by the case), and the aggregate of all ‘other’ delays. These other delays might include the time from source case isolation to swab collection, and the time from source case interview to the infected contact being instructed to isolate. Empirical and modelled distributions for each scenario and contact tracing delay are shown in Figure 3.

For scenarios without case-initiated contact tracing, we estimate the distributions of times from swab collection to case notification and from case notification to case interview from NSW data. We use a modelled distribution for ‘other’ delays with mean and variance of one day since we are not able to estimate these directly from the data.

For scenarios with case-initiated contact tracing, we use the same delays for times from swab collection to notification of the source case, and the ‘other’ delays, but we modify the times from source case notification to interview so that some fraction of these delays (those infected contacts that are identified by cases) are always set to zero, and the remainder are sampled from the estimated distribution of times (some of which are also zero). The zero day delay for cases performing case-initiated contact tracing, reflects an assumption that instructions to notify contacts are sent to cases immediately, and that the case is immediately able to identify close contacts. It is assumed that the time taken from this point to instruct contacts to isolate is the same as for manual contact tracing, and this is included in the same distribution of ‘other’ delays.

We applied the above model to three different delay distributions for NSW:

- Optimal — representing the Optimal TTIQ period in NSW from July 2020–February 2021 when the formal case-initiated contact tracing was not in place and thus we assume that contacts start quarantine after being identified by a source case interview.
- ‘Delta epidemic without case-initiated’ — representing the period in NSW from mid-August 2021– mid-September 2021 when case-initiated contact tracing was in place but we assume that contacts start quarantine only after being identified by a source case interview.
- ‘Delta epidemic with case-initiated’ — as above, except with an assumption that 80% of infected contacts are immediately identified by cases and thus start quarantine on the date of source case notification.

The purpose of the first analysis was to assess how closely our simulation model could reproduce the observed times to isolation from the case data during the Optimal TTIQ period. Once satisfied with our model calibration/performance, the aim of our second and third analyses was to assess whether the formal policy of case-initiated contact tracing reduced delays to isolation. We assessed how closely simulation scenarios where contacts were either immediately identified by cases (scenario with case-initiated contact tracing) or by public health authorities following a case interview (counterfactual scenario without case-initiated contact tracing) match the observed times from infection to isolation from the case data during a period in NSW when a policy of case-initiated contact tracing was in place.

During the ‘Delta epidemic with case-initiated’ scenario, we assumed that 80% of close contacts were readily identifiable by the case (*e*.*g*., household and workplace contacts). This high proportion reflects the fact that stay-at-home restrictions in place during this period minimised the number of social contacts and concentrated infected contacts in household and essential workplace settings where contacts are fewer and more easily identifiable. We would expect the fraction of cases found by case-initiated contact tracing to be smaller under less stringent restrictions.

## RESULTS

Figure 4 displays estimates of times from infection to isolation and TTIQ impact on transmission from case data. We estimate that periods of Optimal TTIQ in reduced transmission by 54% (left panel) and Partial TTIQ resulted in a 42% reduction in transmission (middle panel). Further, in the escalating phase of the Delta epidemic in NSW (mid-August to mid-September 2021), we estimate that TTIQ systems were able to reduced transmission by 40% (right panel). The distribution of times from infection to isolation did not vary significantly over the Optimal TTIQ period, suggesting that the performance of NSW’s TTIQ system was consistent over a range of levels of social restriction.

**Figure 4.**
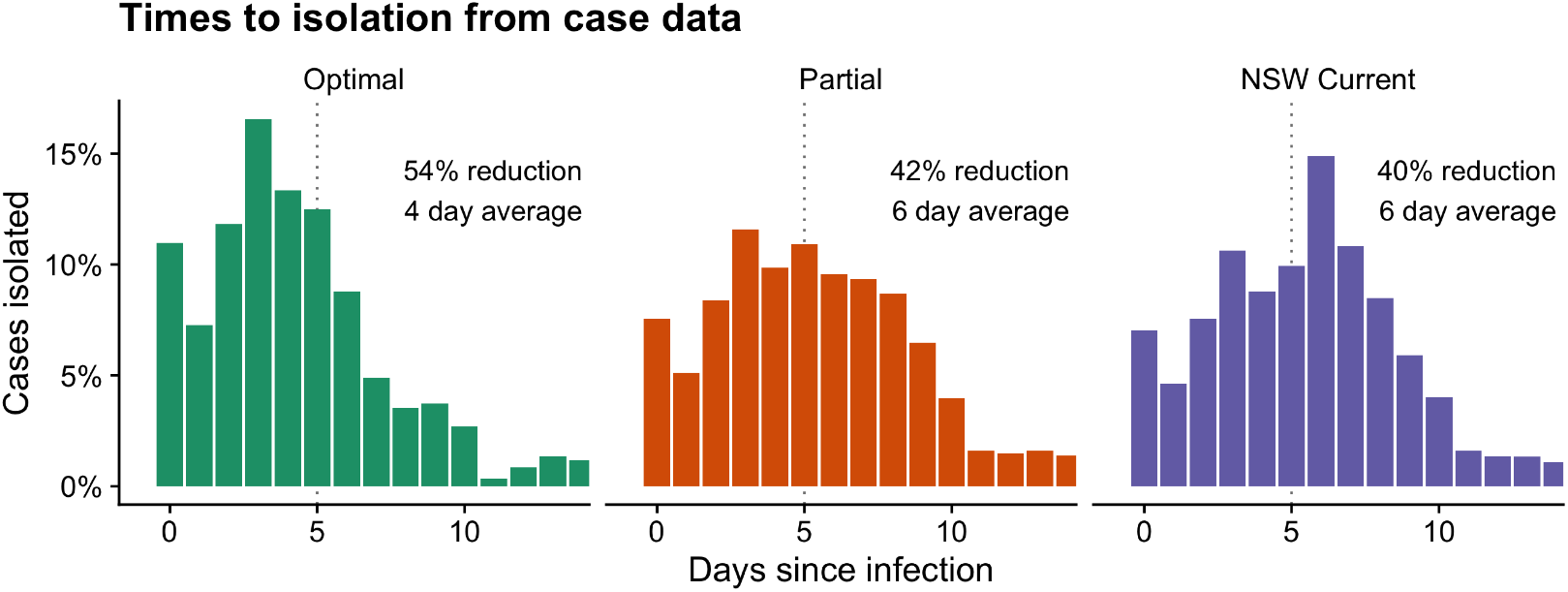
Distribution of delays from infection to isolation, and the resulting percentage reduction in transmission, estimated from case data under three delay scenarios (as outlined in Figure 15). Optimal is times from infection to isolation from NSW case data between July 2020 and January 2021. The distribution of times from infection to isolation for Partial and NSW Delta are extrapolated from Optimal based on delays from symptom onset to notification measured for VIC on 4 August 2020 (Partial) and for NSW on 15 August 2021 (NSW Delta). Dashed vertical lines indicate the time of symptom onset.

Figure 5 displays times from infection to isolation and corresponding TTIQ impact on transmission as predicted by our simulation model under three different delay scenarios. The left panel shows that our simulation model can re-produce the transmission reduction calculated from observed distributions of times from infection to isolation (Optimal TTIQ, 55% reduction). The middle panel shows that with a high level of case-initiated contact tracing, NSW contact tracing delays measured during the escalating phase of the Delta epidemic (August-September 2021) would be able to achieve similar reductions in transmission as for the Partial TTIQ period in VIC 2020 (48% reduction). The right panel shows that the contact tracing delays in NSW during the Delta epidemic are predicted to have resulted in a much smaller reduction in transmission (24% reduction) if case-initiated contact tracing (or other strategies to reduce times to isolation) were not in place. Furthermore, our predicted transmission reduction with case-initiated contact tracing (middle panel, Figure 5) broadly aligns with that estimated directly from NSW case data (right panel, Figure 4). That is, the distribution of times from infection to isolation resulting from our scenario assuming case-initiated contact tracing are closer to the observed distribution of times to isolation from NSW case data, than the counterfactual scenario assuming health authority initiated contact tracing only. This suggests that implementation of a formal case-initiated contact tracing policy was successful in reducing delays to isolation and contributed to transmission reduction during the escalating phase of the Delta epidemic wave in August 2021 when caseloads were thousands per day.

**Figure 5.**
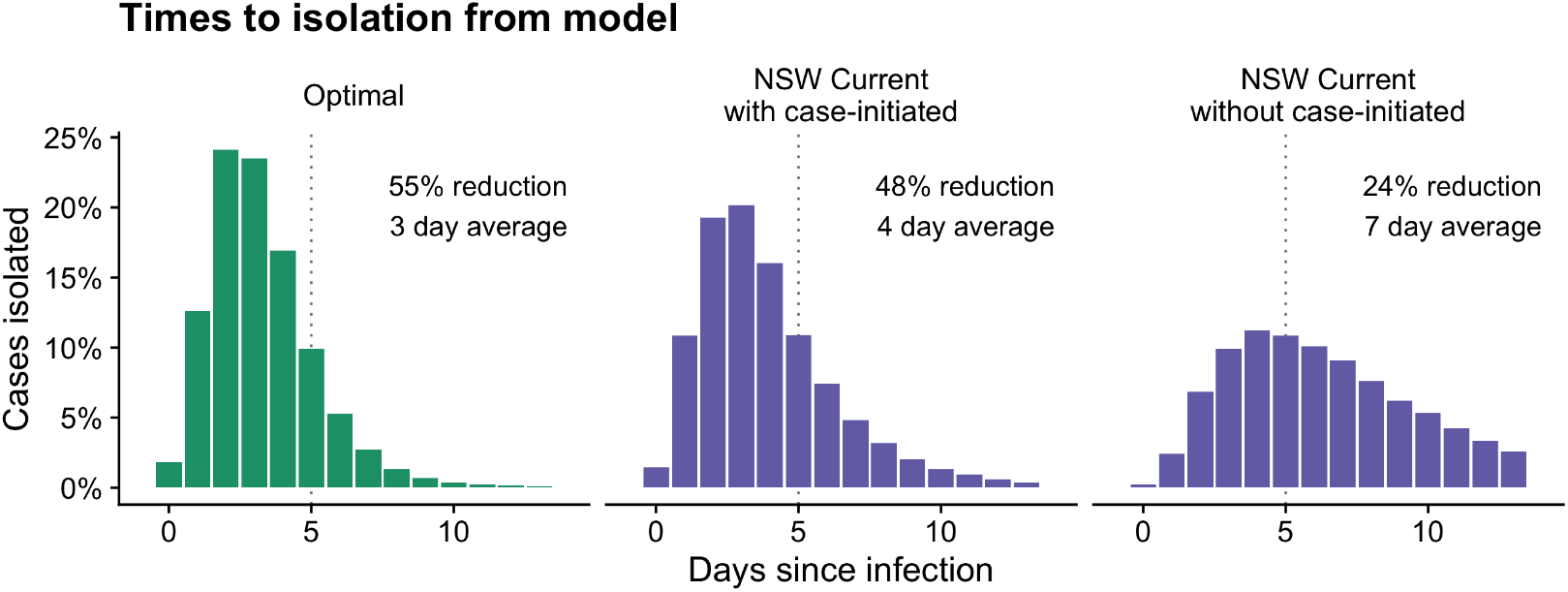
Distribution of delays from infection to isolation, and the resulting percentage reduction in transmission, predicted by our model under three delay scenarios (as outlined in Figure 15). Dashed vertical lines indicate the time of symptom onset.

## DISCUSSION

We assessed the impact of TTIQ on SARS-CoV-2 transmission in Australia during 2020 and 2021, using data from specific locations and times to represent a range of epidemiological and response contexts. We estimate that in a very low prevalence period (tens of cases per day) in the state of NSW, under social restrictions of varying stringency, TTIQ contributed to a 54% reduction in transmission. In a higher prevalence period (hundreds of cases per day) in the state of VIC, under stay-at-home orders, TTIQ contributed to a 42% reduction in transmission. Our analysis also suggests that switching to a case-initiated contact tracing policy in NSW during the 2021 Delta epidemic (thousands of cases per day with a peak of 1,511 cases) contributed to the maintenance of high levels of TTIQ effectiveness under (relatively) high case loads.

These findings must be interpreted in light of the COVID-19 epidemiology and response policy from 2020–21 in these jurisdictions, and our key modelling assumptions. Our analyses assume perfect detection of infections (*i*.*e*., 100% case ascertainment), which we believe is reasonable for Australia’s ‘strong suppression’ era. This was due to the large-scale community testing, intensive contact tracing and epidemiological investigation (including extensive use of sequencing to link cases to clusters), with the explicit aim of identifying all infections in chains of transmission. Test positivity was consistently very low [25], very few cases had no clear source of infection [26], and jurisdictions repeatedly returned to periods of zero case incidence following outbreaks (Figure 1), all suggesting negligible undetected transmission.

In the absence of temporal data on the prevalence of infections in NSW and VIC, such as via random population screening as performed in the United Kingdom [27, 28], we cannot robustly estimate case ascertainment. If case ascertainment were less than 100%, then actual reductions in transmission due to TTIQ would be lower than estimated here, since undetected infections are less likely to isolate. The percentage reduction in transmission is approximately linearly proportional to the fraction of infections detected. For example, if only half of infected people are detected, only half of infections can have any onward transmission events averted due to isolation.

Our analyses assume perfect compliance with isolation and quarantine. That is, all individuals are assumed to stay in isolation/quarantine from the isolation start date (either reported or inferred) for the remainder of their infectious period. Data are not available to estimate levels of isolation compliance, but we expect that it was very high in Australia during the analysis period due to both support to maintain isolation, including welfare checks and counselling, and financial penalties for non-compliance of up to $5,000 enforced through home visits by police/military [29]. Nonetheless, compliance was unlikely to be perfect (encompassing both individuals who partially isolate and those who do not isolate at all), and would have resulted in lower reductions in transmission due to TTIQ than estimated here.

Our estimates of the TTIQ impact on transmission were made for periods when other, synergistic, public health measures were in place, including restrictions on population movement and social gatherings. These measures reduce the pressure on TTIQ systems by both directly contributing to transmission reduction and reducing the per case burden on public health authorities (since on average cases would have fewer contacts per day to trace). Strict stay-at-home measures were in place in VIC during the Partial TTIQ period (August 2020), including: a night-time curfew; one hours’ outdoor exercise allowance; once daily shopping for essential supplies; restrictions on movement more than 5km from a person’s residence; and strict definitions of essential workers and businesses. While population-average rates of daily non-household contacts were significantly reduced during this period, epidemiological assessments suggested that the populations in which transmission was concentrated likely had higher-than-average levels of social contact [20]. Stay-at-home measures were also in place when NSW enacted case-initiated contact tracing (July 2021). When simulating case-initiated contact tracing, we assumed that a high proportion of contacts could be identified by the case, which while likely reasonable under stay-at-home conditions, was unlikely to be possible under lighter social restrictions.

During the Optimal TTIQ period from July 2020–December 2020, NSW effectively controlled a series of localised outbreaks while society remained relatively open (though some restrictions were in place such as social gatherings capped at 20 people). In our previous analysis of this period of the NSW epidemic (using a different modelling approach and data sources including social contact data [20]), we estimated that levels of population-mixing were sufficient to support continued epidemic growth, yet each outbreak was brought under control, which we interpreted as reflecting a strong TTIQ response. The performance of TTIQ in NSW was maintained during a rapidly growing outbreak in mid-December 2020 (resulting from two large super-spreading events), likely supported by stay-at-home orders imposed in the affected locality for three weeks.

The impact of TTIQ on SARS-CoV-2 transmission has been investigated in a number of other settings [30, 31, 9, 15, 32, 33, 34, 35]. These studies use either mathematical models, parameterised to COVID-19 transmission and contact tracing processes [30, 31, 9, 15, 35], or perform empirical analyses of case data [32, 33, 34]. Others have assessed the combined impact of TTIQ and social restrictions by measuring serial intervals over time (time between the symptom onsets of an index case and a secondary case) since the more quickly infected people are identified and isolated, the fewer the opportunities for transmission late in the infectious period, the shorter the serial interval [36]. Disentangling the relative impact of TTIQ measures from other response measures is challenging, partly because interventions are often implemented in rapid succession, but also because much of the case data available to inform such analyses is incomplete or biased in ways that are difficult to measure. In an analysis of the effectiveness of 13 categories of non-pharmaceutical interventions in reducing SARS-CoV-2 transmission using case data from 2020, Liu and colleagues concluded that evidence regarding the effectiveness of testing and contact tracing was inconsistent and inconclusive [32].

Because our study is based on pairs of exposure and isolation times for cases that likely represented a very high proportion of all infections, we are able to directly quantify the impact on transmission of truncating the infectious period of cases via TTIQ measures. However, we are not able to estimate the extent to which these systems may have been assisted by other measures, and therefore our results must be interpreted in context of the other measures in place, as discussed above.

Davis and colleagues performed a mathematical modelling study to estimate potential TTIQ effectiveness in the United Kingdom (UK) in 2020–21 [30]. The model incorporated information on TTIQ systems in the UK (*e*.*g*., test turnaround time, percentage of symptomatic individuals presenting for testing, *etc*.) and used population survey data on isolation compliance. They concluded that well-implemented contact tracing could provide up to a 15% reduction in onward transmission (above and beyond reductions in transmission due to physical distancing behaviour).

It is not surprising that we estimate a much stronger effect of TTIQ in Australia compared to the UK study. The UK study assumed imperfect compliance with isolation (up to 90%), imperfect tracing coverage (up to 80% of contacts reached), and 50% of symptomatic individuals presenting for testing. Further, it does not account for the effects of strategies other than downstream contact tracing to identify contacts, such as the tracing of upstream or secondary contacts, since these were not part of UK test trace isolate procedures at the time (and likely unsustainable under the case burden). In contrast, in Australia contact tracing for COVID-19 from 2020 through to 2021 was not just focused on primary contacts identified by the case, authorities also used location-based contact identification, traced both primary and secondary contacts (when capacity allowed), and routinely performed upstream tracing for cases with no identified source of infection (supported by rapid genomic sequencing) [17]. Location-based contact identification, supported by QR code ‘sign-in’ systems, provided an additional ‘ring’ of contact identification and management. The designation of ‘contacts’ requiring management by these means was much more inclusive than many close contact definitions employed in other countries [9, 37, 38]. Furthermore, the level of presentation by the Australian public for testing was high throughout the analysis period (2020–21), including during periods of low or zero prevalence. During a localised outbreak in December 2020, NSW recorded 69,809 tests in 24 hours (8.7 tests per 1,000 population), reporting seven confirmed cases in that period [25].

In addition to assumptions of perfect case ascertainment and perfect compliance, there are a number of other limitations to the analysis presented here. Our model assumed that in the absence of a formal case-initiated contact tracing policy, infected contacts would not isolate until after being identified by a case interview. However, cases may inform household members and other close contacts of their positive result, and these contacts may choose to self-isolate. The predominance of such self-directed isolation behaviour will likely be difficult to estimate from data, since recorded dates of isolation could represent the date when contacts are instructed to isolate, the date when they began isolating, or the last day in the community, but not all three dates. To simulate case-initiated contact tracing in the escalating phase of the Delta epidemic in NSW, we reduced the time from case notification to notification of their contacts (*i*.*e*., removing the delay case notification to case interview). While our model incorporating these assumptions was able to closely match the observed times to isolation, it is possible that other strategies implemented by health authorities or changes in population behaviour during this period may have also reduced delays to isolation.

We did not examine the effectiveness of TTIQ response in Australia’s six other states and territories, largely due to very low (or zero) case incidence over the analysis period (and hence scarce data available for analysis). However, we expect that the TTIQ systems in these jurisdictions were high-performing, since similarly intensive TTIQ strategies as used in NSW and VIC were employed in response to SARS-CoV-2 incursions. For example, in response to an incursion in November 2020 (fewer than 50 cases in total), the state of South Australia imposed state-wide stay-at-home restrictions and employed intensive tracing of primary and secondary contacts, placing contacts in hotels for the duration of their quarantine period [18].

Our study demonstrates the high impact of TTIQ on SARS-CoV-2 transmission in Australia from 2020–21, prior to the emergence of the Omicron variant in November 2021. High-performing TTIQ systems were both critical to the maintenance of Australia’s strong suppression strategy and directly assisted by the low prevalence environment. Our analysis also highlights the value of surveillance data that enables the evaluation of intervention effectiveness. Direct estimates of TTIQ impact would not have been possible without systematic collection of the dates of likely exposure and isolation of cases by public health response teams in NSW. These data supported both real-time operational decision-making, including local case and contact management, and longer-term strategic response planning, with estimates of TTIQ impact incorporated in a range of modelling analyses supporting Australia’s 2021 national COVID-19 ‘re-opening’ plan [21].

## Data Availability

The analyses performed in this study required access to data from the Australian National Notifiable Disease Surveillance System (NNDSS) and the New South Wales (NSW) Notifiable Conditions Information Management System (NCIMS), which are not able to be shared under the terms of our data access agreements. For access to NNDSS data, a request must be submitted via. For access to NCIMS data, contact the NSW Ministry of Health via https://data.nsw.gov.au/contact.

https://github.com/njtierney/ttiq-simulation

## CODE AVAILABILITY

Code for performing the analyses and generating the figures is available at: https://github.com/njtierney/ttiq-simulation.

## ETHICS STATEMENT

The study was undertaken as urgent public health action to support Australia’s COVID-19 pandemic response. The study used data from the Australian National Notifiable Disease Surveillance System (NNDSS) provided to the Australian Government Department of Health under the National Health Security Agreement for the purposes of national communicable disease surveillance. Data from the NNDSS were supplied after de-identification to the investigator team for the purposes of provision of epidemiological advice to government. Contractual obligations established strict data protection protocols agreed between the University of Melbourne and sub-contractors and the Australian Government Department of Health, with oversight and approval for use in supporting Australia’s pandemic response and for publication provided by the data custodians represented by the Communicable Diseases Network of Australia. The ethics of the use of these data for these purposes, including publication, was agreed by the Department of Health with the Communicable Diseases Network of Australia.

The study also used routinely collected patient administration data from the New South Wales (NSW) Notifiable Conditions Information Management System (NCIMS). Data was supplied by the NSW Ministry of Health, with all records de-identified and securely managed to ensure patient privacy and to ensure the study’s compliance with the National Health and Medical Research Council’s Ethical Considerations in Quality Assurance and Evaluation Activities. The NSW Public Health Act (2010) allows for such release of data to identify and monitor risk factors for diseases and conditions that have a substantial adverse impact on the population and to improve service delivery. Following review, the NSW Ministry of Health determined that this study met that threshold and therefore provided approval for the study to proceed. The project oversight and approval for publication was provided by the NSW Ministry of Health.

## ACKNOWLEDGEMENTS

This work was directly funded by the Australian Government Department of Health Office of Health Protection. Additional support was provided by the National Health and Medical Research Council of Australia through its Centres of Research Excellence (SPECTRUM, GNT1170960) and Investigator Grant Schemes (JMcV Principal Research Fellowship, GNT1117140; FMS Emerging Leader Fellowship, 2021/GNT2010051).

This research was supported by The University of Melbourne’s Research Computing Services and the Petascale Campus Initiative.

This work could not have been completed without the support of the public health network in NSW. The Public Health Response Branch, NSW Health including contact tracing staff as well as operational, policy, epidemiology, surveillance, data quality and data acquisition staff contributed to the TTIQ implementation and data provision. In addition, colleagues in the public health units located within the 15 local health districts across the state all contributed to the implementation of the TTIQ strategy.

